# Community Memory of COVID-19 Infections Post Lockdown as a Surrogate for Incubation Time

**DOI:** 10.1101/2020.04.15.20067058

**Authors:** Meher K. Prakash

**Affiliations:** Theoretical Sciences Unit, Jawaharlal Nehru Center for Advanced Scientific Research, Jakkur, Bangalore 560064; VNIR Biotechnologies Pvt Ltd, Bangalore Bioinnovation Center, Helix Biotech Park, Electronic City Phase I, Bangalore 560100

**Keywords:** Incubation time, COVID-19, Coronavirus, Linear response, Memory function, Lockdown

## Abstract

If the knowledge of the incubation time is helpful in designing non-pharmaceutical interventions such as quarantine measures, can one use the number of cases arising after a lockdown to check (*a*) if our assumptions of the incubation time were correct and (*b*) if the quarantine measures were as successful as they could theoretically be. These are the two questions we raise by studying the number of new cases arising after lockdowns in a few European countries. The analysis which purely relies on the publicly available data of the numbers of new infections, rather than extensive contact tracing of individual patients, suggests a “memory” of the infections in the community with a median of 13.3 days. This distribution of the memory of infections which may even be considered as a surrogate of the incubation time in a perfect lockdown, suggests that even a perfect quarantine of 30 days is only 90% complete.

## Introduction

In designing non-pharmaceutical interventions such as quarantine measures, it is important to know how long an individual can potentially remain infected and infectious.^[1]^ The incubation time of an infectious disease is the duration between the exposure to an infected individual and the appearance of the symptoms.^[2,3]^ The incubation times for several acute respiratory infections have been measured^[4]^ assuming their distributions. ^[5]^ These incubation times range from a couple of days to about 2 weeks.^[4]^

In the studies of the COVID-19 pandemic, incubations lasting 2 to 14 days^[6]^, and reports suggesting up to 19 days^[7]^ or even 24 days^[8]^ have been observed with median incubation times of around 5.1 days^[9]^ to 5.8 days^[10]^. At the heart of the estimation of the incubation period is an efficient contact tracing. The symptomatic individuals history is traced back to the possible day of exposure to infected individuals. As rigorous as this procedure is, by its nature it is not easily scalable as the collecting the exposure histories of hundreds of individuals may not be easy. This problem is even more complicated in the COVID-19 infections, as a very large fraction of the infections have been passed on by asymptomatic individuals. Further, the number of patients studied is also limited, which makes the likelihood of the errors in the “tail regions” of the distribution much higher.

Almost all countries went into a lockdown for several weeks. However, the new cases of infection continue to arise, despite the best quarantine practices many democratic countries could adopt. This raises several question – are estimates of the incubation period based on a few hundred cases reliable, or is there an alternative way to estimate this incubation period even if one did not have the detailed exposure histories? Or is there an alternative estimation one needs to do at the level of the community with the best quarantine practices possible? Are the new cases in a lockdown scenario arising from incubation or because of other factors? We address some of these questions by proposing a new way of estimating the incubation time.

## Results and Discussion

### A new surrogate for incubation time

We propose a new way of estimating the incubation time only from the publicly available epidemiological data of the number of infections. The two advantages of this approach are that the exposure history is not required and in a pandemic one can work with data tens of thousands of infections. In the publicly available data on the number of new infections from different countries, one sees that the number of daily new cases (*I*(*t*)) actually *increase* for a few days (*t*) after the lockdown (*t*=0), before they decline. In an ideal scenario, the new cases after lockdown come from the individuals who were infected in the preceding days and after a certain incubation are becoming symptomatic. Thus the number of new infections on day 1, depends on the asymptomatic ones on day 0 (*A*(0)) and an incubation of 1 day, as well as the ones infected on day −1 and with an incubation of 2 days, and so on. Mathematically, the predicted number of infections *I\**(*t*) on day *t* after the lockdown is

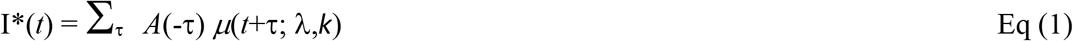

where *μ*(*t*+τ; λ,*k*) represents the distribution of the incubation period which we assume to be a Weibull distribution with parameters λ and *k*, without loss of generality. Convoluting two functions such as in Eq. (1) is a standard approach in the linear response theory. We further assume that on any day prior to the lockdown the number of asymptomatic infections is proportional to the number of reported infections (*A*(-τ) = α.*I*(-τ)). We performed a linear regression analysis between the infections observed after lockdown *I*(*t*) and the predicted ones *I*\*(*t*) for various choices of λ and *k* (Figures 1A,B,C). The values of λ and *k* for which the regression coefficient was the highest was considered the maximum likelihood estimate of the incubation period (Figure 1D).

**Figure 1.**
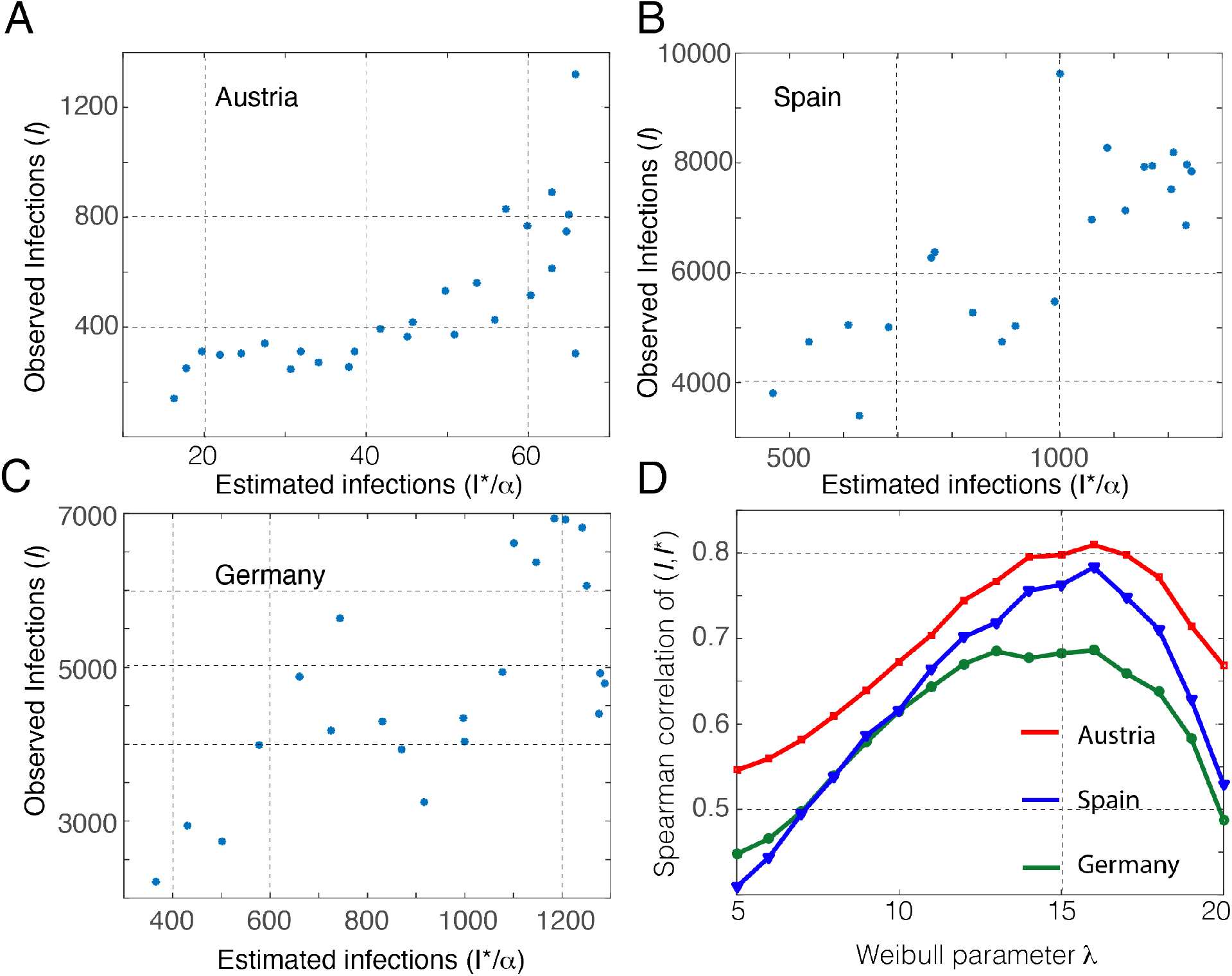
The daily infections observed after lockdown^[11]^ and the predictions made using the infections before lockdown for: ***A***. Austria (lockdown, 16 March 2020) ***B***. Spain (lockdown, 14 March 2020) ***C***. Germany (lockdown, 21 March 2020) are compared. The data until 13 April 2020 was used. The predictions were made as described with λ=16, *k*=2. ***D***. Similar graphs like ***A, B, C*** were generated for λ between 5 to 20 to identify the value that best suits the data. λ=16 optimizes the Weibull distribution for the data studied. *k*=2 was used in this analysis. It was independently optimized in the range *k*=1 to 3.

We find that the incubation time follows a Weibull distribution of *μ*(*t*+τ; λ,k) with parameters λ=16, *k*=2 best helps the infections before lockdown from Austria, Spain and Germany (Figure 1D) to predict the ones after. In working with the epidemiological data, we developed an estimation of the incubation period of the virus in the community, which may in principle be the same as that for the individuals in a perfectly implemented quarantine.

### New cases beyond the incubation

Clearly this incubation is not sufficient to explain the new cases in all countries. If one predicts the new infections post lockdown in Italy, one sees there are more infections than what may be expected purely from a median incubation of 13.3 days (=λ.(ln2)^1/*k*^) for the said distribution (Supplementary Figure 1). However, since there is no reason to believe that the nature of the infections in Italy are different from its neighbor Switzerland (which also has similar incubation time as the countries discussed here), it appears that one should look beyond the incubation to understand the persistent infections.

### Why is the incubation longer?

A Weibull distribution with parameters λ=16, *k*=2 has a cumulative probability that reaches around 0.9 after 30 days. This suggests that the new estimate of the incubation proposed in this work will require a lockdown longer than 30 days for a complete elimination of the asymptomatic cases. The longer incubation time may be a consequence of either an imperfect implementation of the lockdown or a genuinely longer incubation time in individuals that could not be estimated by contact tracing a few hundred individuals. We believe in the latter hypothesis, although in principle the nature of the conclusions do not change regardless of the causative factor. Practically, even if the former is the case, the approach we take is still relevant because several countries have implemented the most stringent measures. In that sense, although physiologically it may slightly differ from the incubation of the individual, this incubation at the community level with the lowest possible societal interactions has its own meaning. In this sense, it is imperative that one uses the incubation period of the infections in the community level into the public health models.

## Conclusions

In summary, we introduced a new way of estimating the incubation time by studying the new infection that arise after measures that restrict the interactions between the individuals in the community to a minimal or non-existent level. The advantages are that it is easy estimate. The estimates we made for three European countries suggest an incubation time with a median of 13.3 days. Models for public health should consider estimates for incubation time in the community, such as what we introduce here, for a better understanding of the differences between new cases arising after the incubation and the genuinely newer ones.

## Data Availability

The analysis was built using the publicly avalable data from the Johns Hopkins University. All the scripts and data used in this work will be made available upon request.

## Supplementary Figure

**Supplementary Figure.**
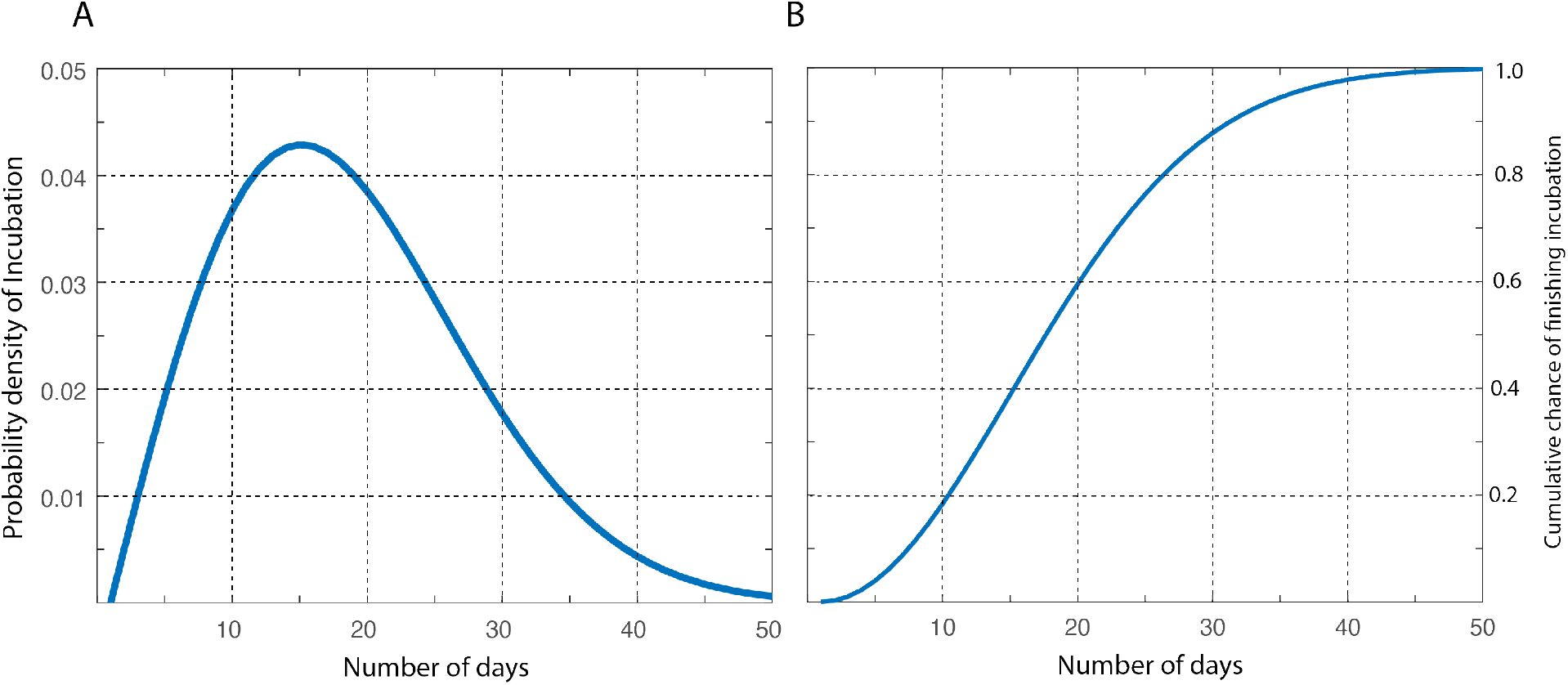
Using the incubation time estimated in Figure 1, μ(t; λ=16, *k*=2) (A) probability density of incubation and (B) cumulative probability that the incubation is finished are plotted. This distribution suggests that even after 30 days, the incubation finishes only with a 90% chance.

